# Blood flow restriction resistance training as an alternative to resistance training alone to improve strength in elderly: a systematic review with meta-analysis

**DOI:** 10.1101/2023.01.19.23284773

**Authors:** André Luiz Silveira Mallmann, Leonardo Peterson dos Santos, Lucas Denardi Doria, Luis Fernando Ferreira, Thiago Rozales Ramis, Luís Henrique Telles da Rosa

**Author notes:** Corresponding author e-mail address, Hospital de Clínicas de Porto Alegre (HCPA)., 2350, Ramiro Barcelos St., 90420-010, Porto Alegre – RS, Brazil. conceptualization; data curation, investigation, methodology, project administration, resources, software, validation, visualization, roles/writing – original draft. data curation, formal analysis, methodology, software, validation. formal analysis, investigation, methodology, software. conceptualization, investigation, methodology, visualization. conceptualization, methodology, supervision, visualization, writing – review & editing. conceptualization, data curation, methodology, supervision, validation, visualization, writing – review & editing.

## Abstract

The purpose of this research was to perform a systematic review with meta-analysis to compare the effects of resistance training with blood flow restriction (BFR) to the effects of non-training (CON) and traditional RT on strength in elderly people. This was a systematic review with meta-analysis of randomized clinical trials (RCTs), published in English, from inception to 2022, conducted using MEDLINE (PubMed), EMBASE, Web of Science and Cochrane Library. The methodological quality was assessed using GRADE protocol. The risk of bias was assessed using RoB2 software. Standardized mean differences (SMD), mean difference, were pooled using a random-effects model. A p < 0.05 was considered statistically significant. Eight RCT’s were included. We found no significant differences in the effects between BFR and RT (SMD = -0.18 [-0.56 to 0.19]; p = 0.34; I^2^ = 12%). Also, evidence from our research shows that the effect of BFR is better than non-training (CON) for strengthening in older adults (SMD = 0.63 [0.24 to 1.01]; p = 0.001; I^2^ = 11%). Our primary findings show that training with BFR may be an alternative methodology of training for the elderly and this training strategy may be interesting for health professionals working with elderly people with low tolerance to high intensity RT.

## INTRODUCTION

The world’s population, according to UN data, could reach 2.1 billion people over the age of 60 by 2050. With the increase in life expectancy, the need arises to find alternative interventions capable of preserving or even improving the health, functionality, and autonomy of individuals of older age groups [1]. Aging generates a sharp decline in the capacity to produce strength, based on the neurological and musculoskeletal systems, a factor that determines autonomy and, consequently, the quality of life of the elderly [2]. It has been discussed the importance of strength for health in people in advanced age groups, since there is an important relationship between this ability and the performance of simple daily tasks for older people [3,4]. The literature has shown that the reduction in the ability to produce strength may be related to several health conditions that negatively influence the quality of life of the elderly, such as an increased risk of limited mobility, increased dependence for activities of daily living (ADL), cognitive decline in older elderly, and an increased risk of mortality from all causes [5-7].

Although the resistance training (RT) is referred to in the scientific literature as an effective strategy for preserving or improving strength [4,8], there are certain situations, like people with joint injury, osteoarthritis and other conditions that make its application not recommendable or difficult to apply due to low tolerance presented by these populations [8-10]. Over the last two decades, a training modality that has stood out in research around the world is resistance training associated with vascular occlusion, also known as Blood Flow Restriction training (BFR), which is the main subject of this review [11-14]. BFR is characterized by using tourniquets or cuffs that create a blockage in blood vessels in one or more members of the human body. Concomitantly, RT exercises are performed with the limbs submitted to vascular occlusion in this training strategy [13].

Studies have demonstrated the efficiency of this method, which has been showing strength gains similar to those generated by high intensity RT in young and elderly populations [11]. However, the main advantage in using this training strategy in populations that are unable to use high intensities in RT, due to joint limitations or due to dysfunctions, is the use of much lower weights to produce a satisfactory strength improvement, as mentioned, since the effect, resulting from BFR, seems to demonstrate better results with lower intensity when compared to BFR associated to high intensity(amount of weight shifted)[13]. Studies show that the adequate intensity for the BFR to present more satisfactory results seems to be approximately between 20 and 50% of 1RM for young populations, values that are determined according to other variables, such as the choice of exercises, volume, and density of the training sessions [13]. Thus, BFR emerges as a viable alternative for a population that suffers from disorders of the neurological and musculoskeletal system, resulting in a lower risk of injury from RT and, therefore, justifying the need for a greater understanding of the efficiency of this intervention [14,15].

Although there are several studies that have been willing to understand the effects of BFR on the capacity to produce strength, there are no systematic reviews that have evaluated the influence of BFR on strength in people over sixty years of age. a systematic review by Centner et al, (2019) sought to compare the effects of BFR to the effects of RT on strength in older people. However, the population included in this research consisted of people over the age of fifty [16]. In this study, we restricted the population to people over the age of sixty, taking into account that this is a population that has very specific needs and limitations, as mentioned as well as a difference from people over fifty years in neuromuscular and functional characteristics that are naturally associated to aging. There is a lack in the literature that needs attention to help health professionals to better understand this intervention. For the reasons mentioned, the aim of this review was to compare the effect of strength training associated to vascular occlusion with traditional resistance training and/or non-training on strength in people older than sixty years.

## METHODS

### Research registration

We conducted this systematic review with meta-analysis in accordance with PRISMA statement (see Supplemental Digital Content 1 that is a table with PRISMA check list) [17]. Search commission of Federal University of Health Sciences from Porto Alegre registration number: 084/2020. PROSPERO registration number: CRD42020220729.

#### EXPERIMENTAL APPROACH TO THE PROBLEM

This was a systematic review with meta-analysis based on a focused question described in a PICO format [18]. We established: Patient/Problem/Population = Elderly (age ≥ 60 years), Intervention = Resistance training with blood flow restriction, Comparison = Resistance training/placebo/non-training, Outcomes = Strength and Study design = Randomized clinical trials.

All studies were screened and assessed for eligibility regarding our inclusion and exclusion criteria, which were based on the PICOS principle (i.e., extracting population, intervention, comparison intervention, outcome measures and study design information).

### Data sources

The electronic databases used were MEDLINE (PubMed), Web of Science, Cochrane Library, and EMBASE in February 2022. We used a comprehensive search strategy tailored to each database. In cases of missing data, authors of selected papers were contacted. When contacted authors did not answer, data were extrapolated from figures, using Image J software, or using available data and mathematics formulas provided by Cochrane’s handbook as presented at section 2.4 [19].

### Search strategy

For identification of relevant studies, a systematic literature search was performed by two blinded researchers (Mallmann, ALS & Doria, LD). Keywords and medical subject headings (MeSH) for the terms “Blood flow restriction”, “Kaatsu”; “Vascular occlusion”, “Strength training”, “Resistance Training”, “Muscle strength”, “Elderly”, “Older” and “Aging” were selected. No filters were used to perform this search. The term OR was used for Union of MeSH terms and “entry terms”, and the term AND was used to attach the terms. The complete string used at PUBMED and adapted for the other databases is fully described at Supplemental Digital Content 2. Study information, including title and abstract, were exported from the databases and stored in a citation manager (Mendeley1, version1.17.9). Before further processing of the studies all duplicates were removed.

## PARTICIPANTS

### Inclusion and exclusion criteria

We included: (1) Participants were older than 60 years, (2) the study design allowed to compare resistance training combined with blood flow restriction to traditional resistance training and/or to a control group (without any intervention or placebo training, such as light stretching), and (3) strength were assessed pre- and post-intervention. No restriction on publication date was imposed.

We excluded: (1) participants received any kind of substance that could interfere on study results, (2) the manuscript was not written in English, Portuguese, or Spanish languages or (3) meta-analysis articles.

Tables with inclusion/exclusion decisions by reviewer and agreements can be accessed at Supplemental Digital Content 3

## PROCEDURES

### Data extraction and assessment of reviewer agreement

First, two researchers (Mallmann, ALS and Doria, LD) screened, independently, all the studies titles, then abstracts and, finally, the full text of the included papers. This process was made in three steps and the reviewers were blinded about the coworker screening. In case of any discrepancy, a third reviewer (Dos Santos, LP) were asked to find an agreement and decide to include or not the paper. The sheets of inclusion/exclusion process for Mallmann, ALS and Doria, LD is available at Supplementary material 2. All data from each study were screened using a bibliographic management program (Mendeley1, version1.17.9).

After screening of the studies, all relevant considered articles were assessed for eligibility based on their full texts. At this stage, we extracted information about (1) population characteristics, (2) primary outcome measures, (3) methods, (4) exercise/interventional characteristics and (5) the main result of the study. When intervention effects were assessed at multiple time points, only the very last time point was considered (as post-training value). When available, data were extracted in the form of delta mean (mean_change_), delta standard deviation (SD_change_), and sample size of the studies to perform the meta-analysis. In case of incomplete raw data availability, we contacted the corresponding author of the manuscript or extrapolated the data from figures, if the authors could not be reached. When the article reported baseline and post-intervention outcomes, however, without mean_change_ and SD_change_, we used the equation (Delta mean = post-training mean– baseline mean) to calculate the delta value. Considering cochrane’s handbook recommendations to calculate the SD_change_, we used the correlation equals zero, since none of the selected papers provided the delta data as mean ± pattern deviation [19]. In order to find SD_change_for selected studies, the following formula was used.

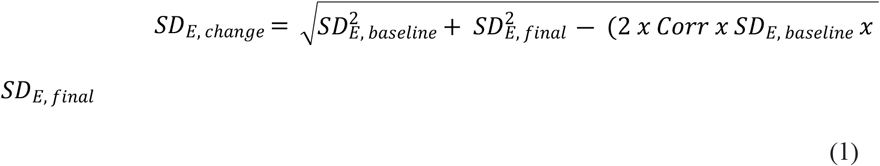

Where Corr is correlation coefficient in the experimental group, SD_E,baseline_is baseline standard deviation in the experimental group, SD_E,final_ is final standard deviation in the experimental group and SD_E,change_ is standard deviation of the changes in the experimental group. When data were presented by interquartile range (IQR), it was decided to transform these data in order to standardize the results of all studies in mean_change_ and SD_change_. The equation used to calculate the mean_change_ is available below [20].

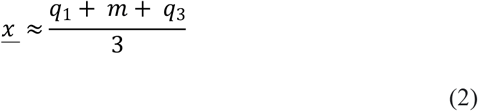

Where q1 is the first quartile, m is the median and q3 is the third quartile. Finally, to find the SD_change_ presented by IQR, we use the calculation available below [20].

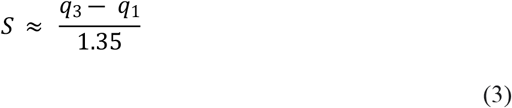

The choice for using these formulas was based on a previous Systematic Review with meta-analysis about effects of BFR training on strength, hypertrophy, and functionality for people with osteoarthritis and rheumatoid arthritis [21].

The extracted data of included studies are sample characteristics (number of participants, age, Body Mass Index (BMI), Duration of the intervention (weeks), frequency (sessions per week), sets, repetitions, interval (seconds), one maximum repetition percentual, blood flow restriction pressure (BFR mmHg) and the delta strength, resulting from interventions).

### Methodological quality assessment

Methodological quality of reports was determined using the Grading of Recommendations Assessment, Development and Evaluation (GRADE). The peer reviewed analysis is available at supplementary material 4. For each of the 7 items of the GRADE scale, two reviewers (Mallmann, ALS and Doria, LD) assessed the studies independently. Disagreements about methodological quality were resolved by a third reviewer (Dos Santos, LP). The GRADE approach considers the risk of bias and the body of evidence to rate the certainty of the evidence into one of four levels:

#### High certainty

We are very confident that the true effect lies close to that of the estimate of the effect.

#### Moderate certainty

We are moderately confident in the effect estimate — the true effect is likely to be close to the estimate of the effect, but there is a possibility that it is substantially different.

#### Low certainty

Our confidence in the effect estimate is limited — the true effect may be substantially different from the estimate of the effect.

#### Very low certainty

We have very little confidence in the effect estimate — the true effect is likely to be substantially different from the estimate of effect.

Studies were included independently of the methodological quality calculated.

### Risk of bias

The risk of bias of the studies was assessed using the risk of bias tool 2.0 (RoB2) from Cochrane [22]. Two authors (Mallmann, ALS and Doria, LD) independently assessed the risk of bias. In the case of disagreement, the subject was discussed with another author (Dos Santos, LP). The evaluators analyzed the randomization process, deviations from intended interventions, missing outcome data, measurement of the outcome, and selection of the reported results. The studies were classified into low, moderate, or high risk of bias.

## STATISTICAL ANALYSES

We conducted a meta-analysis using mean_change_ and SD_change_ from each study. All outcome measures were continuous variables. Two meta-analyses, representing the effects of interventions, were performed: the random-effects model with the mean difference (MD) or standardized mean difference (SMD). MD was performed when studies reported outcomes with the same assessment scale or instrument. When the same outcomes between studies were evaluated but analyzes by different scales or instruments, we performed SMD [19]. The calculation of SMD is represented by dividing the difference in mean outcome between groups by the standard deviation of the result within the groups. The formula between groups within each study used is available below [21,23].

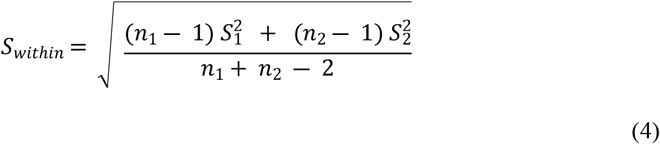

The 95% confidence intervals (CI) were used, and the heterogeneity of the studies included in the meta-analysis was assessed using the inconsistency test (I^2^). Inconsistence was considered as low, moderate or high when values were 25%, 50% and 75% or more respectively [19,24]. The software used for statistical analysis was RevMan (Review Manager 5.4.1, The Cochrane Collaboration, 2020), and we considered significant statistically when P < 0.05.

## RESULTS

### Search strategy

The searches were performed in the five databases from beginning to 2022 and returned 6,956 (380 duplicates) (Fig 1). After removing duplicates, reading the titles, abstracts and full texts, eight studies, between 2013 and 2020, were kept for analysis. Six of them compared BFR training to a non-training group (CON) [25-30] and six of them compared BFR training to a group trained with traditional moderate or high intensity RT [25,27,29,31,32].

**Fig 1.**
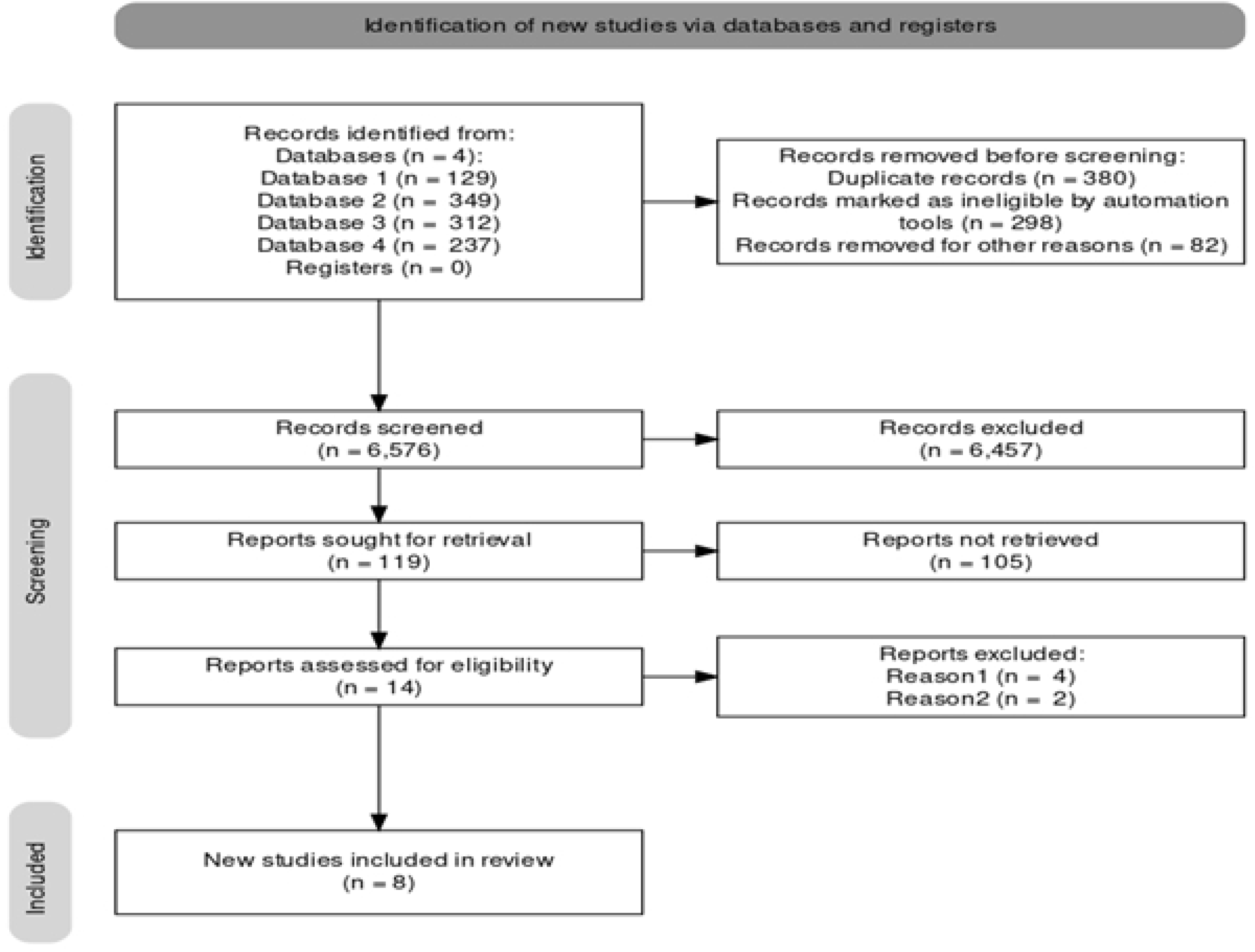
Flow chart including literature search and selection steps following PRISMA statement.

### Meta-analysis data

Fig 2(A) shows the comparison of BFR with RT for selected papers. Only studies with similar methodologies were included in this analysis, minimizing the risk of biasing findings. Studies ere included in our meta-analysis if the assessments of strength were made preferentially with bilateral knee extension (KE) exercise. We also included, for this meta-analysis, papers that evaluated participants with Leg press and unilateral KE exercises. Although, a sensibility analysis, represented by figure 2(B), was made by excluding the study from Letieri et al, 2018 because the training was slightly different from the others for the use of unilateral KE, whereas the other authors compared bilateral KE for both groups. Nevertheless, there were no significative differences between training methodologies, even without considering said study [27].

**Fig 2.**
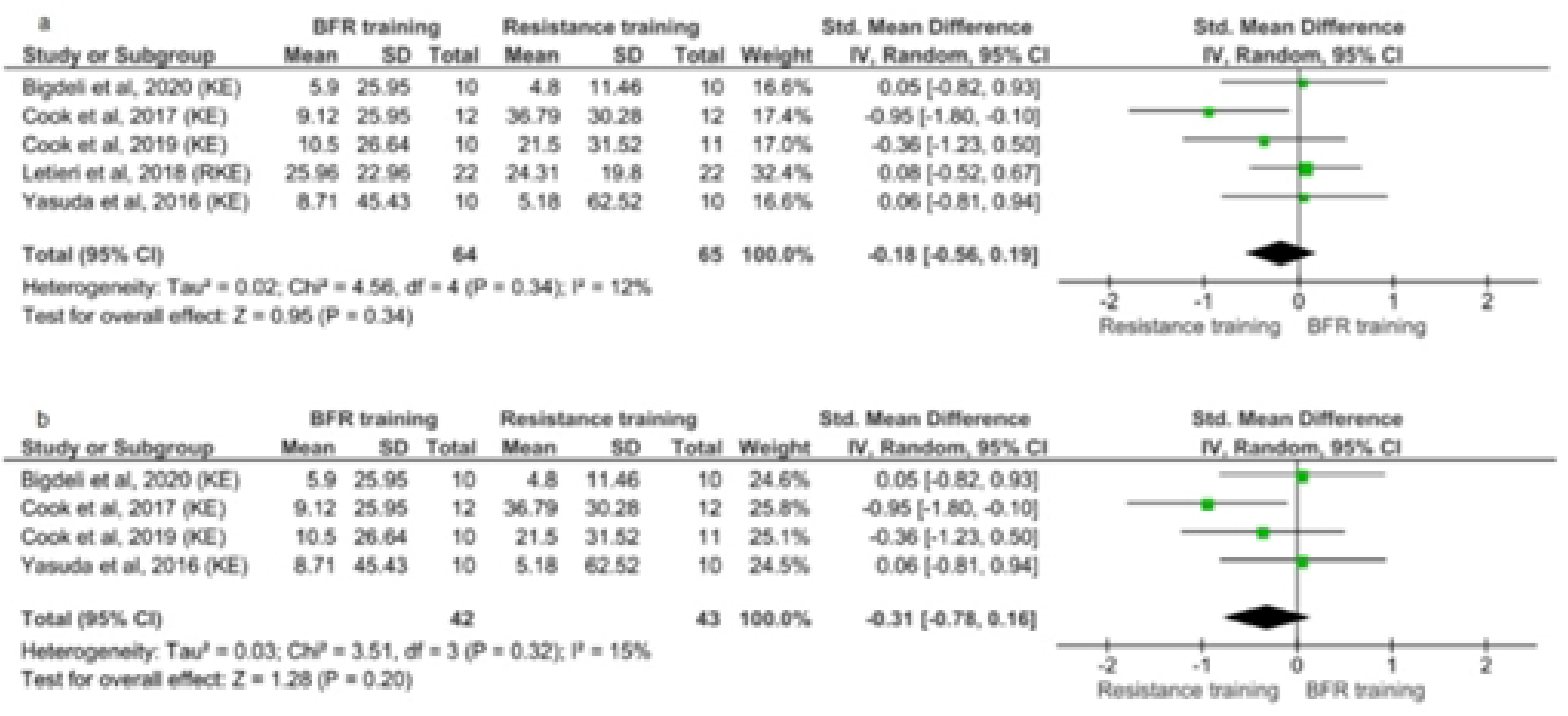
Forest plot graph of the strength of elderly trained with BFR versus Resistance training. A: comparison for all studies with similar methodology including unilateral and bilateral KE; B: sensibility analysis excluding the paper from Letieri et al, 2018 that used unilateral knee extension; *KE:* knee extension; *BFR:* blood flow restriction;*I*^*2*^:inconsistency test (heterogeneity); *SD:* standard deviation; *Std:* Standardized, *95% CI:* confidence interval, *IV:* inverse variance, *Random:* random effects model

Fig 3 shows the comparison between BFR and CON and its sensibility analysis. Figure 3 (A) is the overall comparison between all studies that included groups without any intervention between assessments or applied placebo training (e.g., light stretch training). Figure 3 (B, C and D) illustrates the sensibility analysis we made to minimize the bias of that comparison. Were excluded papers with methodological differences that could interfere on results. First (B) we excluded studies from Letieri et al, 2018 and Vechin et al, 2015 for using unilateral KE and Leg Press (LP), respectively. Second (C) we reintroduced only the study from Vechin et al, 2015 and at least we excluded this one again and reintroduced that from Letieri et al, 2018. Although, the difference between groups remained significative in favor of BFR training.

**Fig 3.**
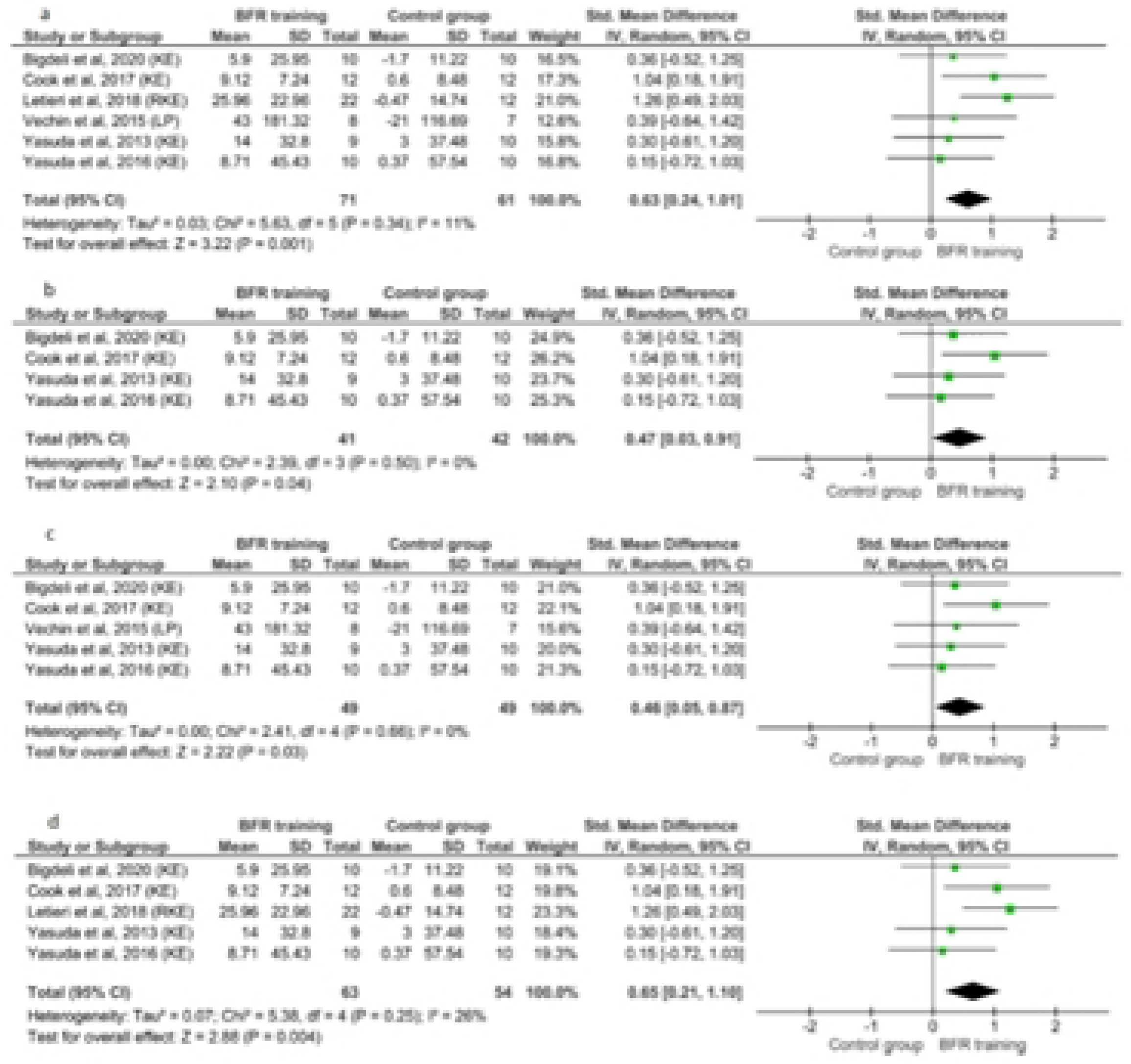
Forest plot graph of the strength of elderly trained with BFR versus Control group (without training or placebo training).A: comparison for all studies with similar methodology including LP, unilateral and bilateral KE; B: sensibility analysis excluding the papers that used unilateral KE and LP; C: sensibility analysis excluding only the study from Letieri et al, 2018 that used unilateral KE; D: sensibility analysis excluding only the article from Vechin et al, 2015 that used LP; *KE:* knee extension; *BFR:* blood flow restriction; *I*^*2*^:inconsistency test (heterogeneity); *SD:* standard deviation; *Std:* Standardized, *95% CI:* confidence interval, *IV:* inverse variance, *Random:* random effects model.

### Risk of Bias

All selected studies were analyzed for their risk of bias according to the GRADE Approach tool, by Cochrane Collaboration. The data presented were analyzed using RoB 2 software.

Among the analyzed fields, shown in Figure 5, the greatest risk of bias was found in the random sequence generation field, with one study that did not present such variable (12%); at the field about measurement of the outcome, most studies (75%) were classified as “some concerns” for risk of bias, because the methods for assess the outcomes were not the very best way to do that. All selected studies were included in the systematic review, independently of quality assessment results.

For more details about risk of bias, see figs 4 and 5 (for complete peer reviewed risk of bias see the figure at Supplemental Digital Content 4).

**Fig 4.**
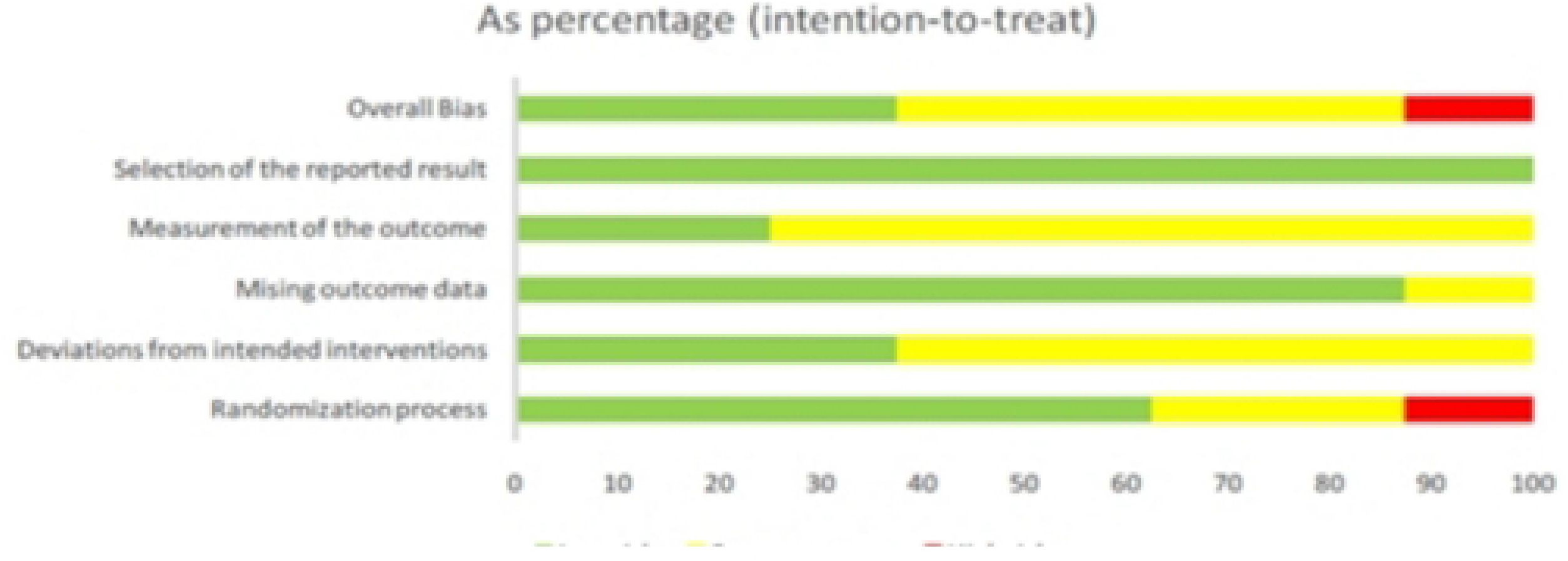
Risk of bias graph considering all studies pooled.

**Fig 5.**
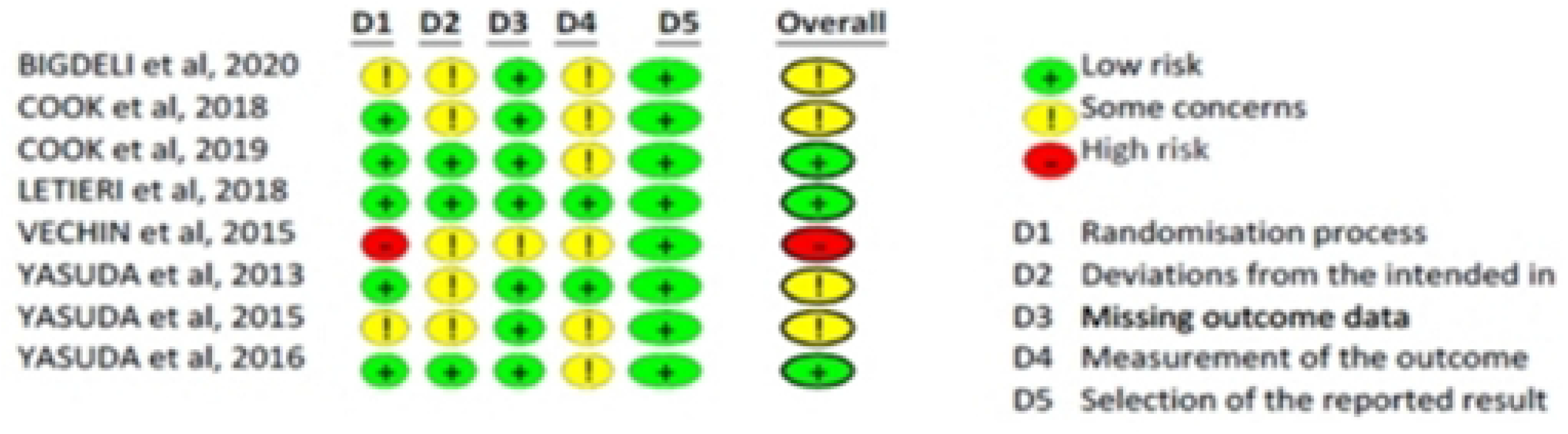
Risk of bias for individual studies and according to the different criteria assessed.

### Quality assessment

Overall studies quality assessment show that the evidence present low quality, in accordance with GRADE tool. The lowest quality shown is for risk of bias domain since a study presented high risk of bias for randomization process. For complete data, see table at Supplemental Digital Content 5 with each reviewer quality assessment.

### Studies Data

In table 1 the characteristics of sample are presented. It is possible to notice that are homogeneity between groups in each study in age and body mass index (BMI). The mean age across studies was 69,2±5,4, and the mean BMI 26,2±2,8. The total sample analyzed was 232 older (≥ 60 years old) men (76) and women.

**Table 1.**
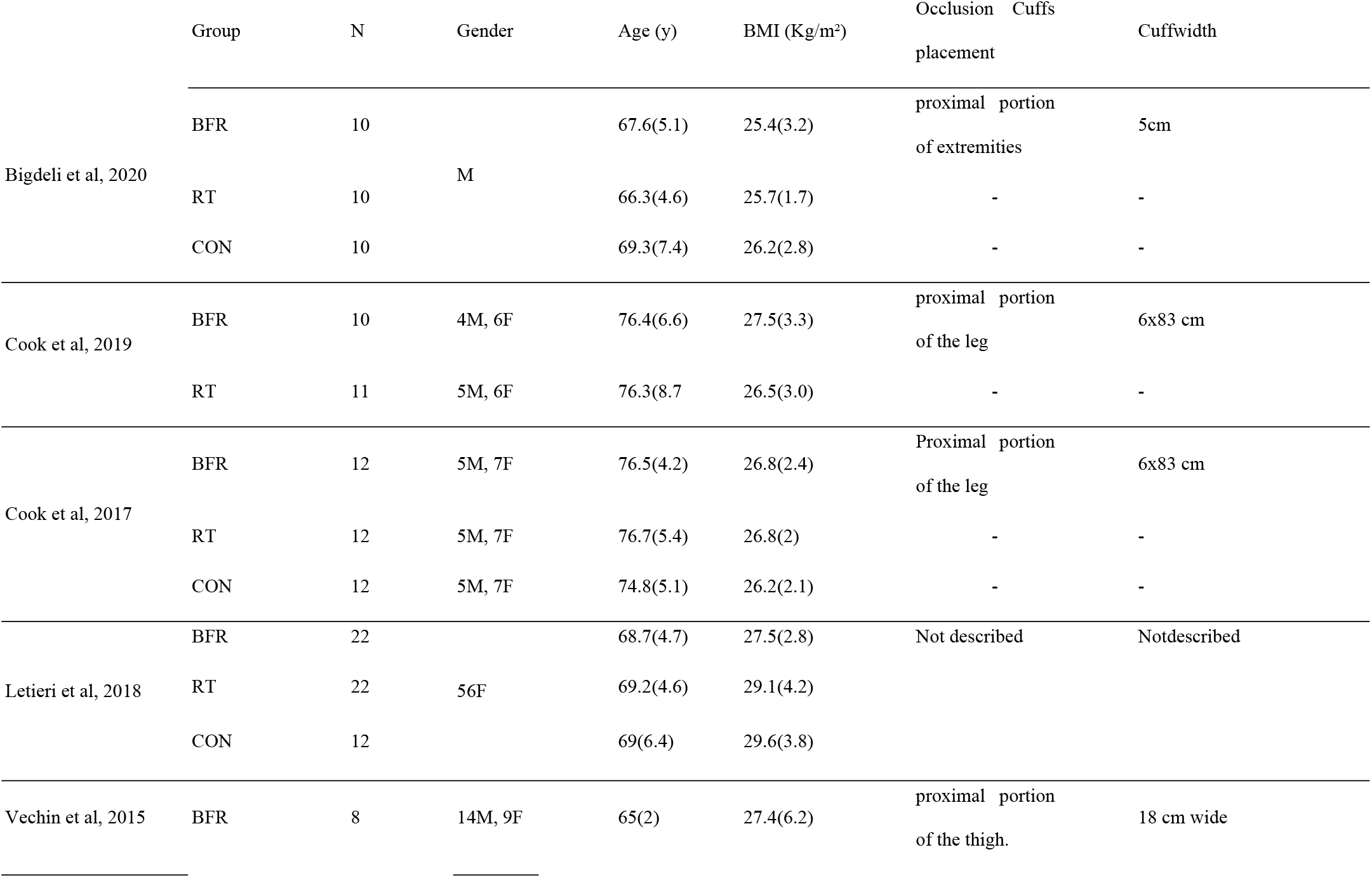
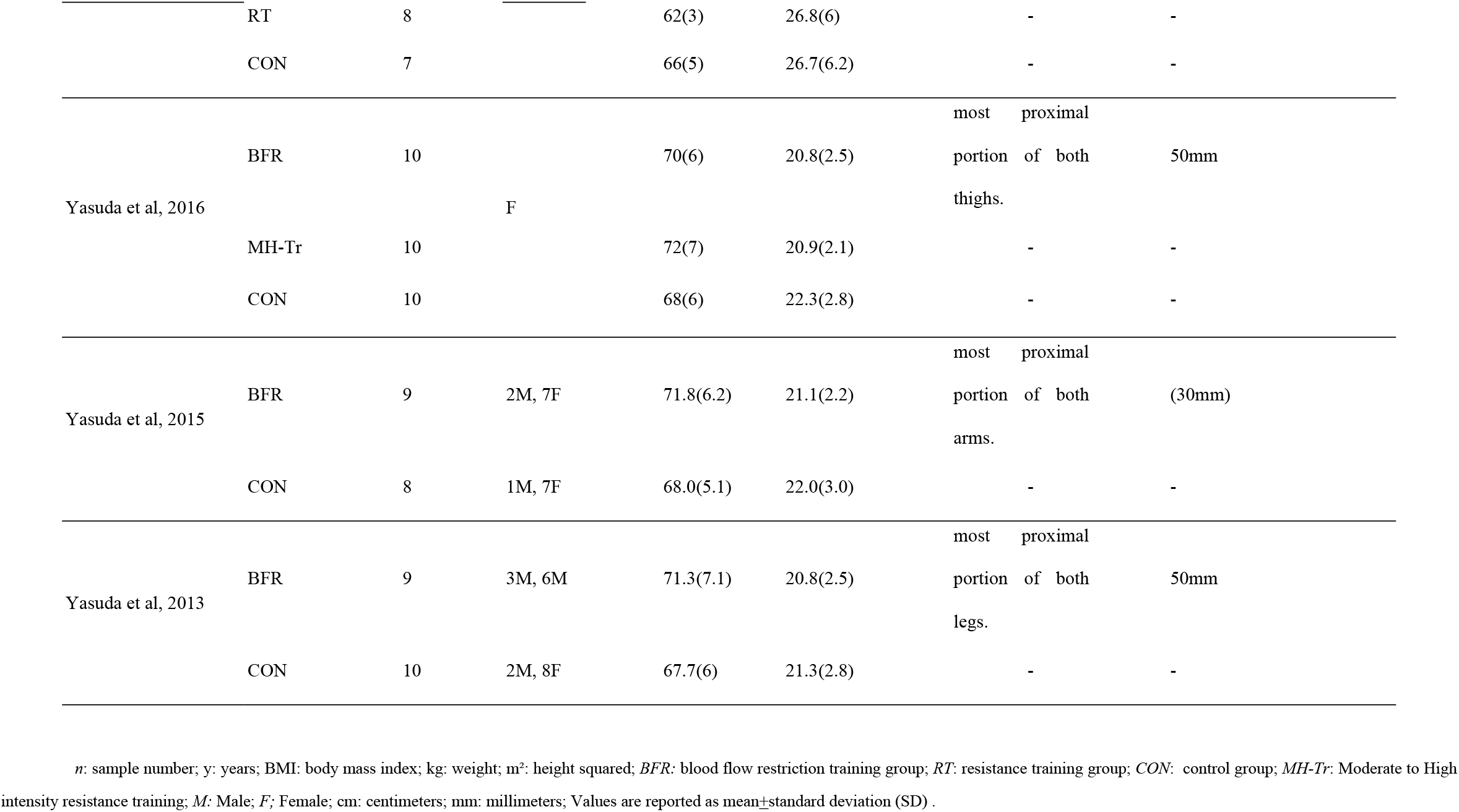
Participant’s characteristics at baseline

Table 2 shows that there is no pattern in the duration of the training period (6 to 16 weeks), in the number of repetitions (from 10 to failure) or in the BFR pressure used (from 71 to 270 mmHg) for the included papers. However, it is possible to observe that the number of sets (2 to 4) and the interval between them (30 to 60 seconds) followed the ACSM recommendation for resistance training [4]. It is also possible to see that for resistance training using the BFR resource, the percentages of a maximum repetition (1RM) were lower (20-30% 1RM) than those of traditional resistance models (70-80% 1RM).

**Table 2.**
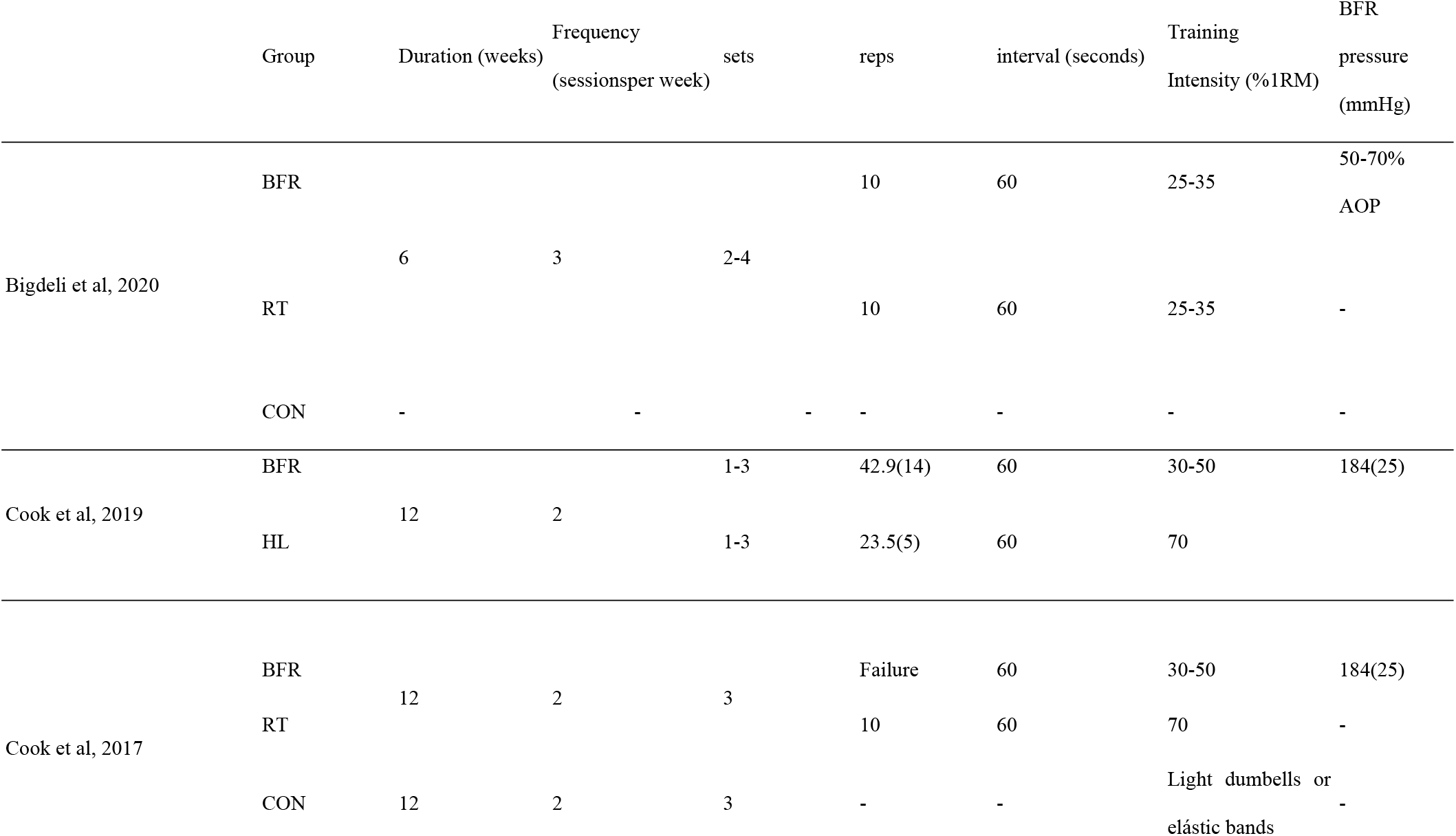
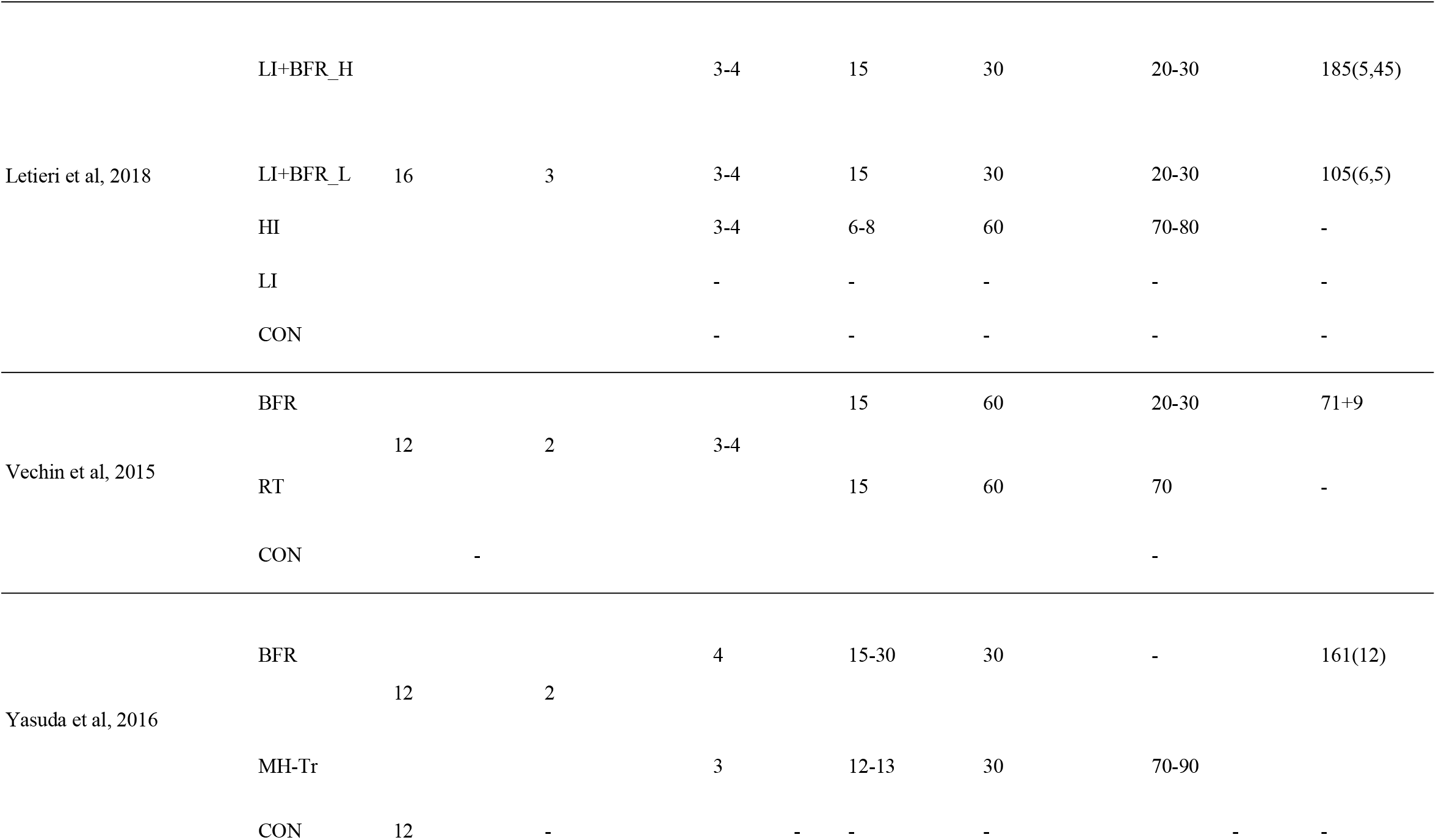
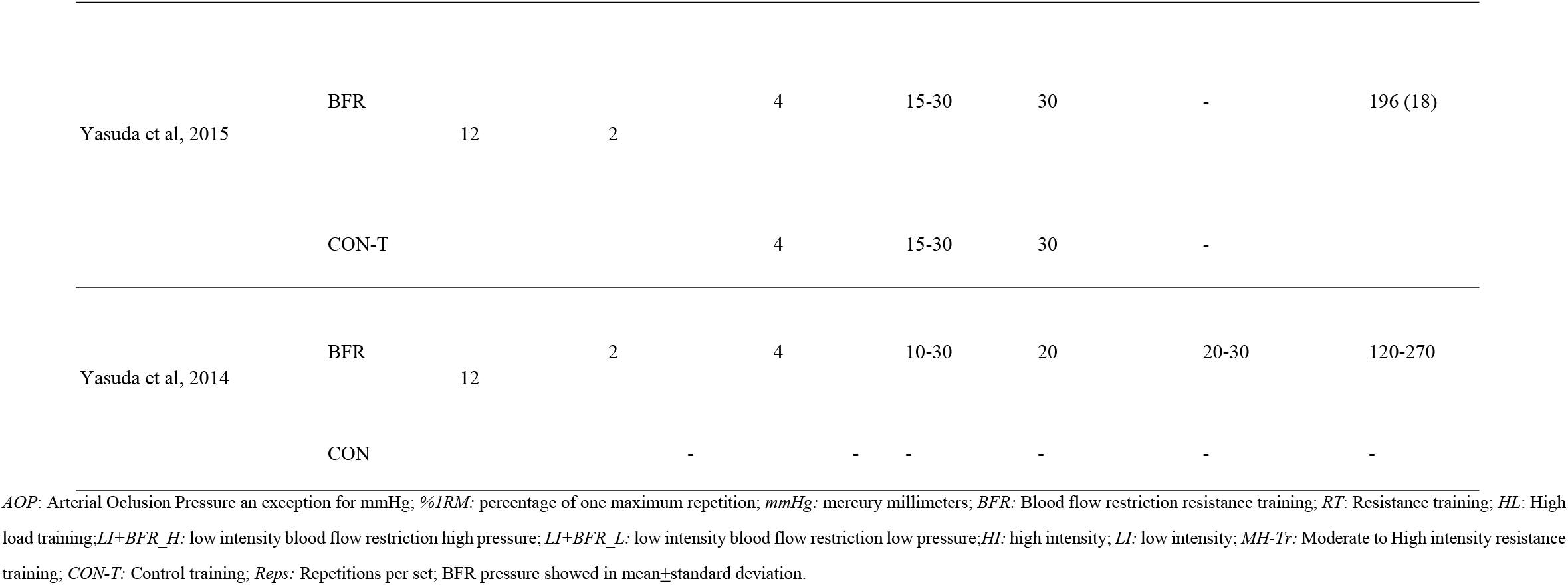
Characterization of the training in the studies resulting from the search in the databases.

Short training protocols describing for each study are presented at Supplemental Digital Content 6

## DISCUSSION

It is important to note that this was the first systematic review to consider previous papers with active and non-active people aged at sixty or more years trained with BFR and compared to RT alone, as well as to people that did not train. Our main finding is the absence of statistically significant difference between elderly people trained either with BFR or RT which justifies the applicability of BFR for people that presents any kind of intolerance to traditional RT. Also, it is important to observe the lack of standardizing on RCT’s methods, specially talking about training protocols and outcome mesures.

### Blood flow restriction vs Resistance training alone

Our results show, by meta-analysis (Figure 2), that there are no statistically significant differences between groups trained with BFR or RT alone and these findings are in consistency with previous literature [33].

In some cases, BFR showed bigger strength improvements than RT alone (Table 3), while in other studies the results from RT were better than BFR for strength improvement and these cases must be analyzed. Study from Letieri *et al*, (2018) show results favorable to BFR, when compared to RT and the study from Bigdeli *et al*., (2020) show results favorable to BFR only for leg extension exercise [25, 27]. In the other hand, in the studies from Cook *et al*., (2017) and from Vechin *et al*. (2015), RT was more effective than BFR for strength gains [26,28]. Differences between studies that could explain why BFR is better in some papers, as the study from Letieri *et al*, (2018), remains on AOP, training intensity (1RM%), number of repetitions per set, also known as training volume, weekly frequency, between sets intervals [4].

**Table 3.**
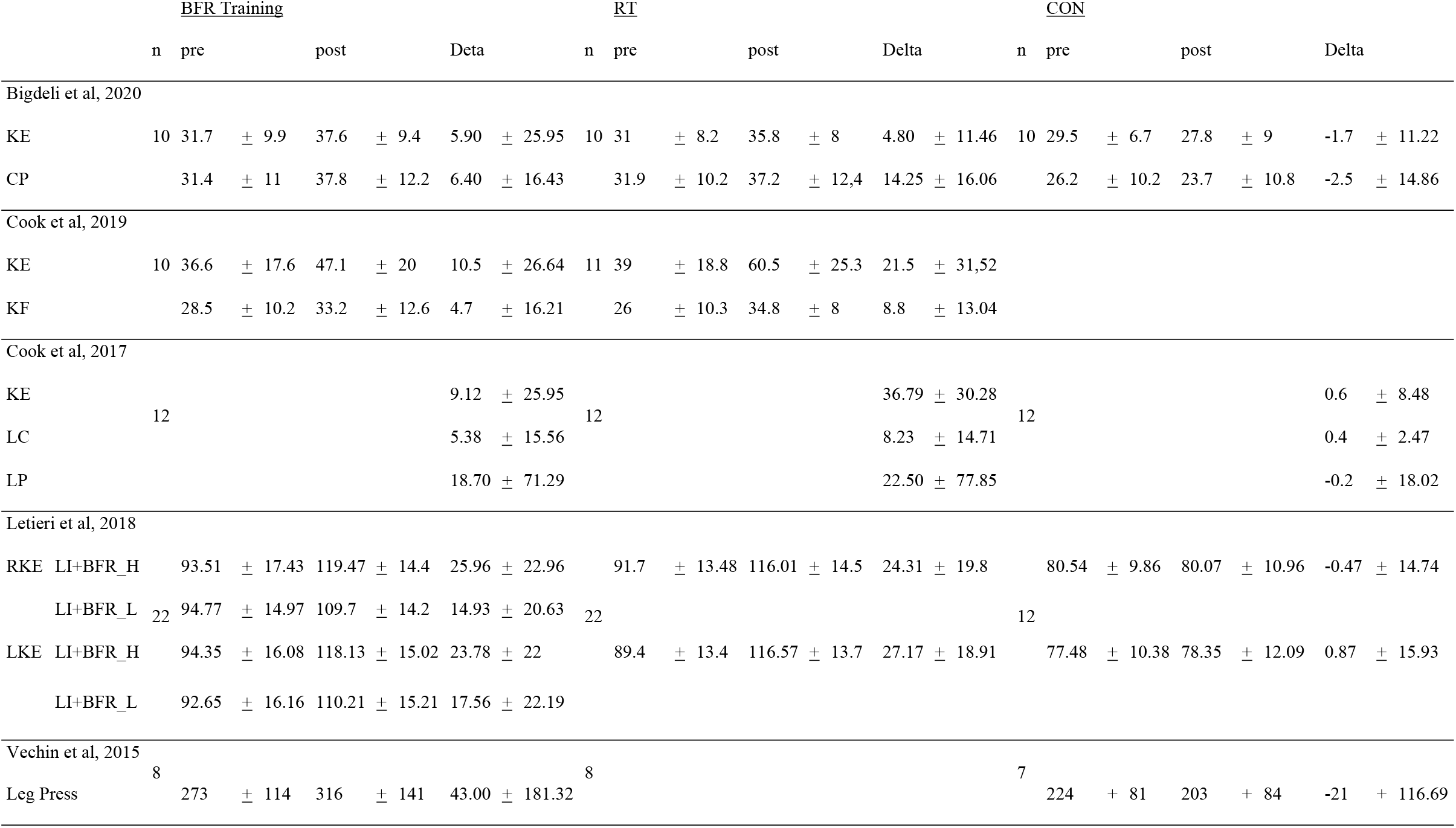
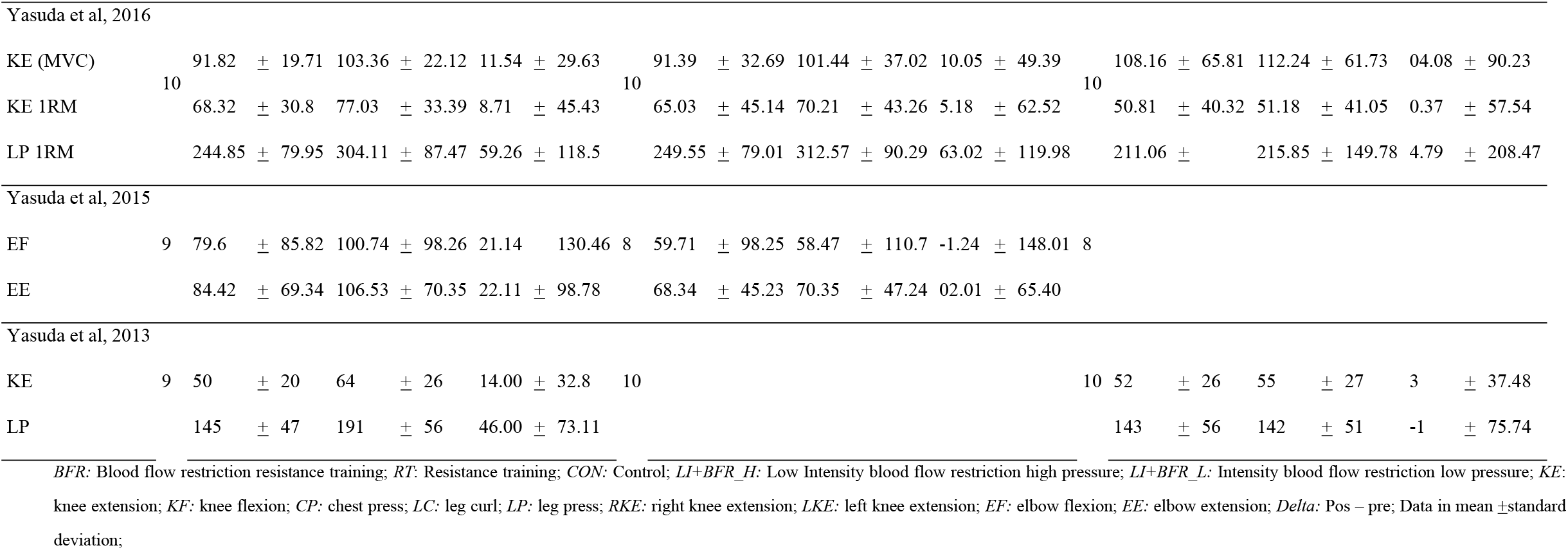
Summary of strength results selected studies

First, Letieri *et al*. (2018) show the best results in favor of BFR and, not coincidently, this is the study with biggest duration, between all selected papers for this review [27]. In line with that, the study from Loenneke *et al*., (2012) show that BFR present better results for strength improvement, with a bigger effect size, when intervention has longer duration, with ten or more weeks from baseline to post-intervention assessments, and this fact could explain our findings at the studies from Bigdeli *et al*., (2020), Cook *et al*., (2017) and Vechin*et al*., (2015), because of the short duration of this studies [25, 26, 28, 33]. It is important to note that the study from Bigdeli *et al*., (2020) was conducted for only 6 weeks, while the studies from Cook *et al*., (2017) and Vechin *et al*., (2015) lasted for 12 weeks, that is, only 2 weeks more than the study from Loenneke *et al*., (2012) found as a time cut point for better strength improvements resulting from BFR training [25, 26, 28, 33]. Another note that deserves our attention is the fact that most of studies about BFR training lasted from 4 to 12 weeks of training, including all kinds of population and this is a reason to difficult a better understanding about this training modality and its impacts on strength due to its short durations.

Although BFR presents less neural drive, our findings show that there is no difference in strength gains between this methodology and RT. A systematic review with meta-analysis from Centner and Benedikt, (2020) found inconclusive results comparing muscle activation between low load BFR training (LL-BFR) and HL-RT, but most of the studies included in this paper lasted for less than 10 weeks, that is the time that, as seen, when BFR starts to show bigger improvement on muscle strength compared to RT [34].

A study from 2021 [35] compared two groups with a repeated measures cross-over design intervention, performed in two sessions of isometric exercises of the knee extensors on two separate days in random and counterbalanced order. The low load group (LL) trained with 20% of the maximum voluntary contraction (MVC) and the low intensity group with blood flow restriction (LL-BFR) trained at 20% of the (MVC), but with 50% of its arterial occlusion pressure (AOP), which was determined in a personalized way before the beginning of the intervention. The findings of this study suggests that BFR is better than traditional RT with low loads for strength improvements and this is an interesting line of thinking, keeping in mind that people with older ages may not be able to accomplish sets of RT with optimal loads for strength gains [35].

We hypothesized that, due to the typical physical limitations of the population within this age, the BFR training could generate gains similar or even superior to the RT since, theoretically, this population would not be able to displace the same percentages of 1RM as a young population. Therefore, they would end up training with low intensity resistance training (LL-RT), a technique that seems to be less efficient, according to the study by Hughes et al., (2017), than BFR training for strength gains [14]. In this 2017 review, the authors sought studies that compared BFR to RT for strength gains in patients with clinical musculoskeletal condition. The research strategy included studies dealt with patients with osteoarthritis of the knees (n=3), ligament injuries (n=3), sporadic inclusion body myositis (n=1) and older adults susceptible to sarcopenia (n=13), corroborating our hypothesis that the older population, which, as stated, has a greater inability to perform training with higher intensities [14].

Although our hypothesis considered that in some cases this population, over 60 years of age, could not perform strength exercises with high loads, our search strategy, which considered medically stable elderly individuals, returned studies that compared BFR training to high intensity RT. And the results of the comparison, performed in this review (Figure 2), suggests that strength gains may be similar to moderate and high intensity RT.

If the neural drive, as mentioned, is smaller with the application of BFR training than with RT, it is important to try to understand how BFR brings strength improvements similar to RT. The literature has shown the physiological mechanisms that could explain these gains and, thus, enabling new hypotheses for future research with greater direction regarding BFR training to control the loss of strength in the elderly [35,36]. Studies show that there appears to be a strong relationship between the metabolic response, arising from BFR training, and gains, both in strength and muscle hypertrophy. The study by Loenneke et al., (2010) was the first to clarifies that the main mechanisms related to gains arising from BFR are apparently the accumulation of metabolites, such as blood lactate, plasma lactate and muscle cell lactate, in addition to being related to activation of fast twitch muscle fibers (FT), even when training intensity is low, and last but not least, gains appear to be related to increased expression of mammalian target of rampamycin (m-TOR) [36].

### Blood flow restriction vs non-training

A meta-analysis comparing a BFR group with a control group was performed. The result of this analysis suggests that BFR training can be an effective alternative for strength gain in people over 60 years, since we found a significative difference between groups in favor of the BFR training one (Figure 3).

This direction has been consistently shown in the literature. A study from Centner *et al*., (2019) compared BFR combined with collagen hydrolysate (BFR+CH) against BFR combined with placebo (BFR+PLA) and to a third group that only consumed the protein supplementation (CH). Both groups with BFR training show improvements in strength capacity while CH group demonstrated a decrease in strength gains, suggesting that BFR is better than non-training to strength capacity, even when participants have protein supplementation [35].

Keeping in mind that there are some elderly people who is not able to perform resistance exercises with high loads, specially following ACSM’s [4] orientations for muscle strength and hypertrophy, due to joint disorders, pain, lack of proprioception and other common conditions of this population, it is important to find therapies or training strategies capable of guaranteeing satisfactory results on the ability to produce muscle strength, insofar as this is a functional capacity that may reduce the risk of falls and mortality as well as increase or maintain the quality of life for elderly who needs to rise from chairs, beds, pick up some objects from the ground and other daily activities that are important for self-autonomy [37, 38].

Our results show that BFR can be an alternative to physiotherapists and other health professionals who needs to find a tolerable training strategy to work with patients with joint injury, pain or any disorder that makes RT not recommendable. This is the main finding of our review because, since muscle strength can be improved with the application of the BFR and this, in turn, requires a much lower intensity than the RT, it can be assumed that this modality would become making exercises with weights more tolerable for people with osteoarticular, neurological and/or musculoskeletal disorders.

### Blood flow restriction parameters

A secondary finding of our study was to compare different protocols of BFR and its effects on strength in elderly. Although, we could not perform any kind of statistical analysis due to heterogeneity of selected papers for BFR protocols, as well as the small number of studies about BFR and its effects on strength for this specific population, making it difficult to carry out subgroups analysis.

It should be observed that the volume (sets x repetitions) and frequency found at the primary studies selected for this review are similar to the guidelines provided by ACSM, except for Yasuda *et al*., (2013) that studied sets up to 30 repetitions [30]. We could not compare different training volumes, intensities and rest intervals. In the same line, it must be noted that there were no comparisons between different occlusion protocols. The only study that made it was the one from Letieri *et al*., (2018) that compared two different levels of pressure applied on individuals trained with BFR [27]. Although, it was not possible to elicit the best protocol with just one study. A systematic review from Lixandrão *et al*., (2018) did this comparison, with a different meta-analysis for each of the BFR characteristics (i.e., cuff width, absolute occlusion pressure, test specificity and occlusion pressure prescription method) and found results in favor of high load resistance training (HL-RT) for all the comparisons. Although, it must be observed the population of said paper, which was composed of young people [34]. Our group sought to perform the same comparison, but with the elderly population, in order to determine whether musculoskeletal deterioration, inherent to advancing age, could interfere with the strength gains arising from BFR training.

A systematic review from 2012 compared several variables of the BFR training, in order to ascertain their influence on the results obtained with this modality. The authors’ first analysis demonstrates that BFR training could be better utilized when associated with low-intensity resistance exercise than when associated with high-intensity. The researchers compared different occlusion pressures and, according to the survey data, there was no significant difference between groups using higher pressures and groups with lower pressures in strength gains and hypertrophy. Thus, it could be suggested that the pressure used for training may perhaps be much lower than that practiced in other studies [33].

By comparing these recommendations to the data obtained from our research, we can draw some conclusions. Regarding training intensity, all studies selected for this review evaluated BFR training associated with similar intensities, ranging between 20 and 35% of 1RM, with the exception of the study from Cook et al., (2017), which used between 30 and 50% of 1RM and, when compared with the traditional RT, it proved to be less effective than this one for increasing strength, corroborating the research carried out in 2012 [26, 33]. As for the occlusion pressure, there are some inconsistencies between our findings and those of that paper. The article by Letieri et al, (2018) obtained greater strength gains using BFR training with both high and low occlusion pressure when compared to traditional RT [27]. On the other hand, the study by Vechin *et al*. (2015) was the one that obtained the worst outcomes from the BFR compared to the RT, although it used the lowest occlusion pressure of all selected studies [28]. Another important training variable is the weekly frequency that is, in most of selected studies composed by 2 sessions for week, with the exception of two papers and, casually, these works found a tendency of higher strength improvements for BFR groups than the RT [25, 27]. A previous systematic review with similar population also found papers with 2 to 3 weeks of duration and, although the authors have not statistically analyzed this variable, results did not show trends to any duration variance as a predictor of better results for strength improvement [39]. It is important to understand the influence of time under ischemic conditions. From selected studies, four training protocols [28-30, 32] made participants remain with inflated cuffs during exercise sets and intervals with an approximate time under ischemic condition of 11 minutes, while three training protocols [26, 27, 31] maintained participants under ischemia during exercises and intervals, but deflated during exercises transitions with a approximate time under pressure of 5 minutes per exercise. Last but not least, one training protocol [25] deflated cuffs between sets and between exercises. Comparing our findings with previous literature [40, 41], we find no pattern for these training variables that may have some influence on training results. This inconsistency suggests future research in order to investigate, in greater depth, the influence of each of the training variables associated with the BFR and its effects for strength improvement.

A potential limitation for this systematic review is the heterogeneity of BFR training and assessments protocols. In some cases, we needed to perform analysis with different exercises (e.g. Leg press and Knee Extension). New papers should be conducted with similar protocols, making it viable to other researchers to compare the same exercises and bring new insights to scientific literature.

In the same line, we could not compare BFR parameters, such as AOP, cuff positioning, time under occlusion, high or low intensity with BFR and others because of the wide range of protocols in a few studies. More research must be made with different protocols and, at the same time, similar protocols, so that systematic reviews can be performed comparing subgroups with different BFR training parameters.

## PRACTICAL APPLICATIONS

Our findings suggests that BFR can be an alternative methodology of training for the elderly who are not able to perform RT with recommended intensity for strength improvement, as meta-analysis demonstrates that there are no significant differences between BFR and RT and also that there is significant difference between BFR and non-training or placebo training (CON).

It was not possible to compare BFR parameters, such as AOP, intensity, volume, density and others. New research should be conducted with the aim to compare different BFR protocols and a better description of methodology, in order to make possible a meta-analysis comparing, for example, occlusion pressures, higher or lower intensities, volumes and densities associated with BFR. Furthermore, it would be interesting for researchers to adopt a pattern regarding the choice of exercises, so that it is possible to compare the BFR and RT using data from multiple studies, making feasible a meta-analysis with these two types of training as comparators.

## Data Availability

All relevant data are within the manuscript and its Supporting Information files.

## ACKNOWLEDGEMENTS

There were no fundings of any kind for this research. We would like to thank Dr. Rafaela Cavalheiro do Espírito Santo, who contributed to the realization of this study with considerations that we applied in the final elaboration of this manuscript.

## DECLARATIONS

### CONFLICT OF INTEREST

André Luiz Silveira Mallmann, Leonardo Peterson dos Santos, Lucas Denardi Doria, Luís Fernando Ferreira, Thiago Rozales Ramis and Luís Henrique Telles da Rosa declare that they have no conflict of interest relevant to the content of this review.

### FUNDING

This research did not receive any specific grant from funding agencies in the public, commercial, or not-for-profit sectors.

### DATA AVAILABILITY STATEMENT

All raw data supporting this systematic review are from previously reported studies, which have been cited. Additional processed data that support the findings of the current review are available from the corresponding author upon request.

### ETHICS APPROVAL

The research was submitted for the Research Committee from Federal University of Health Sciences from Porto Alegre, Brazil with the register number 23103.205151/2020-51. Because it is a review study, it is not necessary to submit for ethics committees.

## Notes

### Competing Interest Statement

The authors have declared no competing interest.

### Funding Statement

The author(s) received no specific funding for this work.

